# National estimates of critical care capacity in 54 African countries

**DOI:** 10.1101/2020.05.13.20100727

**Authors:** Jessica Craig, Erta Kalanxhi, Stephanie Hauck

## Abstract

The COVID-19 pandemic is an emerging threat across the African continent where critical care capacity is underdeveloped or unknown. In this paper, we describe national critical care capacity including number of ICU beds, number of ventilators, and number of physician and non-physician anesthesia providers for 54 African countries. Data was compiled from a variety of resources including World Bank databases, local and international news media, government reports, local healthcare workers, and published scientific literature. Overall, data on number of physicians, hospital beds, and ICU beds were available for over 90% of countries. Data on number of ventilators, number of physician anesthesia providers (PAP) and non-PAP were available for 46 (85%), 47 (87%) and 37 (69%) countries, respectively. Across all 54 countries included in the analysis, there was an average of 3.10 ICU beds and 0.97 ventilators per 100,000 people, and an average of 2.42 total (physician and non-physician) anesthesia providers per 100,000 people. The purpose of this analysis was to fill in knowledge gaps around current critical care capacity across the African continent and to inform national, regional, and international pandemic preparation and response efforts.

## Introduction

Since SARS-CoV-2 first appeared in late 2019, there have been over 5 million cumulative confirmed COVID-19 cases and over 300,000 deaths reported globally.^1^ Many countries have implemented airport closures, curfews, lockdowns, and other social distancing measures to reduce COVID-19 transmission and to prevent health facilities from being overwhelmed by demand for hospital care, intensive care unit (ICU) beds, and ventilators needed to treat severe infections.^2^ Despite these interventions, many well-equipped countries have faced shortages in health equipment and trained personnel.^3,4^

The first confirmed COVID-19 case in Africa occurred in Egypt on February 14, 2020.^1^ Thus far, African countries have reported lower disease incidence with just over 85,000 confirmed COVID-19 cases and 2,308 deaths across the continent as of 27 May 2020.^1^ However, infectious disease surveillance and reporting infrastructure remains highly underdeveloped in many African countries, and COVID-19 testing is limited given the shortage of human resources and appropriate laboratory facilities.^5,6^ In addition, projections of COVID-19 case burden predict that most African countries will experience an uptick in total and severe COVID-19 infections in the next one to three months.^7^

Across Africa, critical care capacity is far below international norms and public health officials have suggested there is a severe lack of ICU beds and ventilators.^8,9,10^ According to a COVID-19 Readiness Survey conducted by the World Health Organization (WHO) in March 2020, an estimated 9 ICU beds are available per 1 million people across the continent.^6^ However, self-reported information from 34 out of the 47 WHO member countries gave a largely incomplete picture of the current situation with regards to the region’s critical care capacity.

To better understand critical care capacity across the continent, we compiled data on number of ICU beds, number of ventilators, and number of physician anesthesia providers (PAP) and non-PAP, among other datapoints, for 54 African countries. This data is intended to inform and assist policy makers and public health officials at the national, regional, and international levels in equipping and preparing African countries to tackle the COVID-19 pandemic.

## Methods

National critical care capacity datapoints relevant to COVID-19 treatment included in the database and subsequent analysis were number of ICU beds, number of ventilators, and number of PAP and non-PAP. Other demographic, economic, and health systems indicators such as number of physicians and hospital beds per 1,000 people were also included. The estimated numbers of ICU beds and ventilators were obtained from published government reports or statements, published scientific literature, reports or statements from aide and other non-governmental organizations, local and international media (in all major continental languages), and in-country informants including government or public health officials and other local researchers and healthcare workers (Appendix 1, 2). Where possible, we cross-checked ICU bed and ventilator data with multiple sources.

The number of PAP and non-PAP was obtained from the World Federation of Societies of Anaesthesiologists Global Anesthesia Workforce Survey.^11,12^

National demographic and economic information for the most recent year for which data was available were obtained from existing databases. Gross domestic product (GDP) at purchasing power parity (PPP) per capita in current international dollar for each country was obtained from the World Bank.^13^ Population data and hospital beds per 1,000 people, and physicians per 1,000 people were obtained from the World Bank’s World Development Indicators database.^14^

Regional sub-groupings of African countries followed those of the United Nations Statistics Division and do not represent official endorsement or geopolitical position.^16^ Disputed and dependent territories were excluded.

For comparisons across countries and regions, we translated available count data and data reported per 1,000 people into rate data reported per 100,000 people.

## Results

### Data Availability

Data availability is summarized in Table 1, and a complete index of data availability is provided in Appendices 1 and 2. Data on GDP PPP per capita, population, hospital beds per 100,000 people, and physicians per 100,000 people were available for over 90% of the 54 African countries.

**Table 1.** Summary of Data Availability

| Variable | Year(s) | Num. Countries with Data out of 54 (%) |
| --- | --- | --- |
| GDP at PPP per capita in current international dollar | 2018 except for Djibouti (2011), Eritrea (2011), and South Sudan (2014) | 53 (98%)<br>Data not available for Somalia. |
| Income group classification | 2018 | 54 (100%) |
| Population | 2018 except for Eritrea (2011) | 54 (100%) |
| Hospital beds per 100,000 | 2004 to 2015 | 52 (96%)<br>Data not available for Eswatini and South Sudan. |
| Physicians per 100,000 | 2014 to 2017 except for Eritrea (2004), Cameroon (2011), Congo (2011), Comoros (2012), and DRC (2013) | 50 (93%)<br>Data not available for Lesotho, Namibia, Sierra Leone, and South Sudan. |
| Number of ICU beds per 100,000 | 2015 to 2020 | 49 (91%)<br>Data not available for Benin, Comoros, Equatorial Guinea, Madagascar, and Mozambique. |
| Number of ventilators per 100,000 | 2017 to 2020 | 46 (85%)<br>Data not available for Benin, Comoros, Congo, Lesotho, Malawi, Mauritius, Seychelles, and Tanzania. |
| PAP per 100,000 | 2015 to 2016 | 47 (87%)<br>Data not available for Central African Republic, Comoros, Equatorial Guinea, Eswatini, Liberia, Sao Tome and Principe, and Somalia. |
| Non-PAP per 100,000 | 2015 to 2016 | 37 (69%)<br>Data not available for Cabo Verde, Central African Republic, Comoros, Congo, Djibouti, Equatorial Guinea, Eritrea, Eswatini, Gabon, Guinea-Bissau, Lesotho, Liberia, Mauritius, Sao Tome and Principe, Seychelles, Somalia, and South Africa. |

Local and international news media were the major sources for data on number of ICU beds and ventilators. Data on number of ICU beds were available for 49 (91%) countries and on number of ventilators for 46 (85%) countries. Data on physician anesthesia providers (PAP) and nonphysician providers (non-PAP) were available for 47 (87%) and 37 (69%) countries, respectively.

It was not possible to discern equipment and human resources capacity at public versus private health facilities or in rural versus urban settings. In addition, we were unable to separately estimate equipment and human resources available for pediatric versus adult patient populations. Therefore, numbers presented here represent total equipment and human resources availability across country and patient segments.

### Critical Care Capacity

Critical care capacity data segregated by income and region is summarized in Table 2, and a complete listing of data is available by country in Appendices 1 and 2.

**Table 2.**
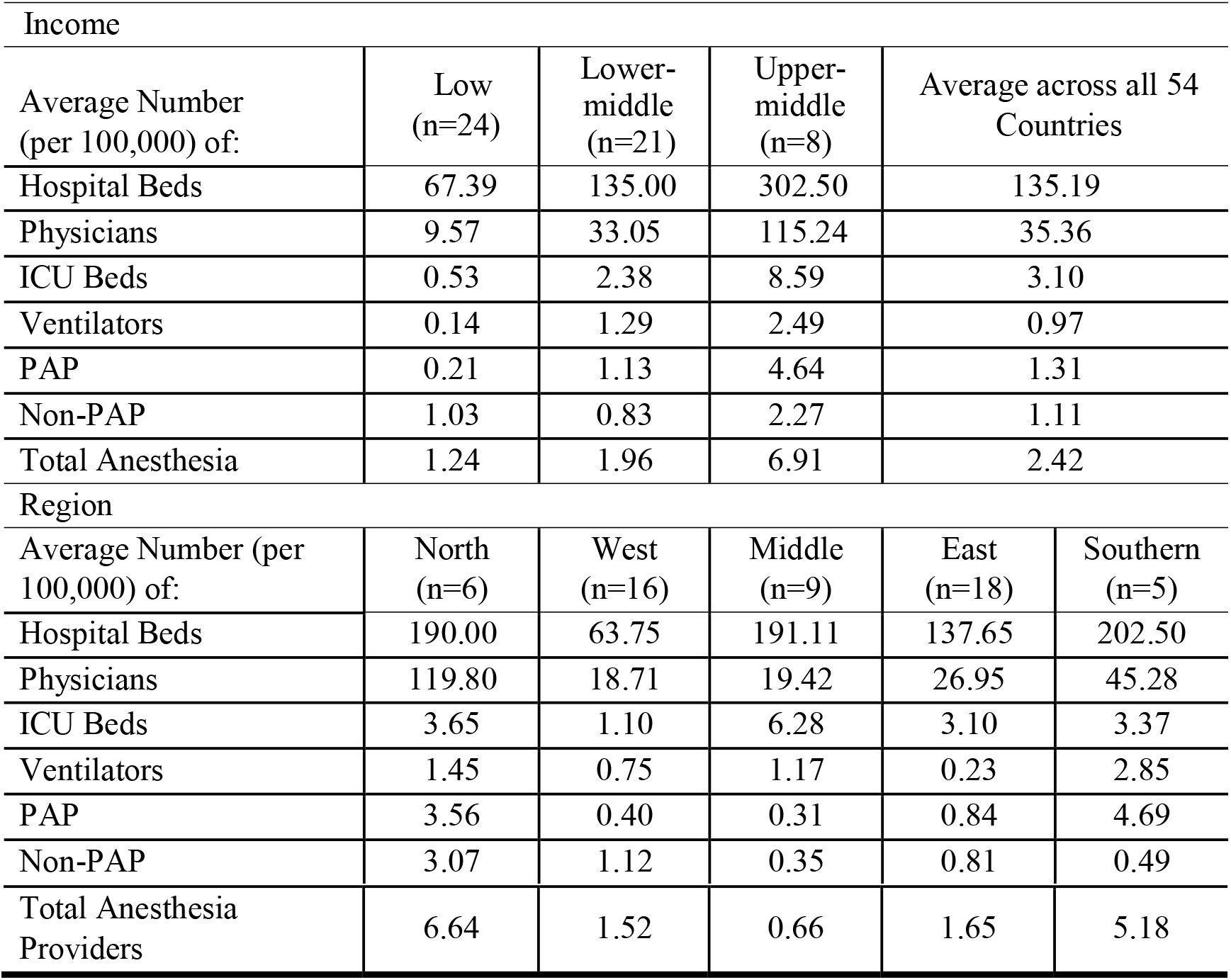
Summary of Critical Care Capacity by Income Group and Region

Across the continent, there were an average of 135.19 hospital beds and 35.36 physicians per 100,000 people ranging from 67.39 beds and 9.57 physicians per 100,000 people in low-income countries to 302.50 beds and 115.24 physicians in upper middle-income countries (Table 2). The average number of hospital beds per 100,000 was highest in Southern Africa and lowest in West Africa while the average number of physicians per 100,000 was highest in North Africa and lowest in West and Middle Africa.

Across all 54 countries included in the analysis, there was an average of 3.10 ICU beds and 0.97 ventilators per 100,000 people. The average number of ICU beds per 100,000 people ranged from 0.53 in low-income countries to 8.59 in upper-middle countries and 33.07 in Seychelles, the sole high-income country included in this analysis. The average number of ventilators per 100,000 people ranged from 0.14 in low-income countries to 2.49 in upper-middle income countries. The average number of ICU beds was lowest in West Africa with only 1.10 ICU bed per 100,000 people, and the average number of ventilators was lowest in East Africa with only 0.23 ventilators per 100,000 people.

Overall, there was an average of 2.42 total (physician and non-physician) anesthesia providers per 100,000 people ranging from 1.24 and 0.66 in low-income countries and in the Middle African region, respectively, to 6.91 and 6.64 providers per 100,000 in upper middle-income countries and the North Africa region, respectively.

## Discussion

Overall, the availability of hospital beds, physicians, ICU beds, ventilators, and anesthesia providers in 54 African countries is far below the capacities of other countries where the demand from COVID-19 has exceeded existing resources.^16,17^ As expected, there is particularly limited critical care capacity in low and lower middle-income African countries. For comparison, in the US, Italy, Germany, and China, there are between 280 and 1,200 total hospital beds and between 240 and 710 ICU beds per 100,000 people.^16,17^

For most countries included in this analysis, there was a lack of verified data available from published scientific papers and reports, or from government Ministries of Health, or other equivalent national agencies. Where possible, we attempted to cross-check our data with multiple sources. In addition, for several countries, we were unable to identify various data points. Despite these limitations, this database on African critical care capacity is the most comprehensive available to our knowledge, and alongside COVID-19 case burden projections, may be useful in guiding and informing national, regional, and continental outbreak preparedness and response.

## Data Availability

All data is publicly available or available upon request from the corresponding author.

## Appendix

**Appendix 1.** Demographic, Economic, and Health Workforce Data by Country with Year of Data*.

| Country | Region | GDP per capita, PPP (current international \$) | Year | Income Group (2018) | Population | Year | Number of Hospital Beds per 1,000 | Year | Physicians per 1,000 | Year | Physician anesthesia provider (2015/2016) | Non-physician anesthesia provider (2015/2016) |
| --- | --- | --- | --- | --- | --- | --- | --- | --- | --- | --- | --- | --- |
| Algeria | North | 15,481.79 | 2018 | Upper middle | 42,228,429 | 2018 | 1.9 | 2015 | 1.8 | 2016 | 1550 | 3500 |
| Angola | Middle | 6,452.36 | 2018 | Lower middle | 30,809,762 | 2018 | 0.8 | 2005 | 0.2 | 2017 | 60 | 210 |
| Benin | West | 2,424.76 | 2018 | Low | 11,485,048 | 2018 | 0.5 | 2010 | 0.2 | 2016 | 61 | 200 |
| Botswana | South | 18,615.79 | 2018 | Upper middle | 2,254,000 | 2018 | 1.8 | 2010 | 0.4 | 2016 | 18 | 0 |
| Burkina Faso | West | 1,984.88 | 2018 | Low | 19,751,535 | 2018 | 0.4 | 2010 | 0.1 | 2016 | 51 | 450 |
| Burundi | East | 744.18 | 2018 | Low | 11,175,378 | 2018 | 0.8 | 2014 | 0.1 | 2016 | 6 | 328 |
| Cabo Verde | West | 7,454.06 | 2018 | Lower middle | 543,767 | 2018 | 2.1 | 2010 | 0.8 | 2015 | 11 | 0 |
| Cameroon | Middle | 3,785.08 | 2018 | Lower middle | 25,216,237 | 2018 | 1.3 | 2010 | 0.1 | 2011 | 83 | 24 |
| Central African Republic | Middle | 859.94 | 2018 | Low | 4,666,377 | 2018 | 1 | 2011 | 0.1 | 2015 | -- | -- |
| Chad | Middle | 1,968.26 | 2018 | Low | 15,477,751 | 2018 | 0.4 | 2005 | 0.0 | 2016 | 1 | 22 |
| Comoros | East | 2,913.40 | 2018 | Lower middle | 832,322 | 2018 | 2.2 | 2010 | 0.2 | 2012 | -- | -- |
| Congo | Middle | 5,662.06 | 2018 | Lower middle | 5,244,363 | 2018 | 1.6 | 2005 | 0.1 | 2011 | 9 | -- |
| Côte d'Ivoire | West | 4,207.09 | 2018 | Lower middle | 25,069,229 | 2018 | 0.4 | 2006 | 0.2 | 2014 | 167 | 304 |
| Democratic Republic of the Congo | Middle | 932.17 | 2018 | Low | 84,068,091 | 2018 | 0.8 | 2006 | 0.1 | 2013 | 98 | 1003 |
| Djibouti | East | 2,744.30 | **2011** | Lower middle | 958,920 | 2018 | 1.4 | 2014 | 0.2 | 2014 | 9 | -- |
| Egypt | North | 12,412.31 | 2018 | Lower middle | 98,423,595 | 2018 | 1.6 | 2014 | 0.8 | 2017 | 5500 | 0 |
| Equatorial Guinea | Middle | 22,743.82 | 2018 | Upper middle | 1,308,974 | 2018 | 2.1 | 2010 | 0.4 | 2017 | -- | -- |
| Eritrea | East | 2,103.00 | **2011** | Low | 3,214,000 | **2011** | 0.7 | 2011 | 0.1 | 2004 | 56 | -- |
| Eswatini | South | 10,637.80 | 2018 | Lower middle | 1,136,000 | 2018 | -- | -- | 0.1 | 2016 | -- | -- |
| Ethiopia | East | 2,022.14 | 2018 | Low | 109,224,559 | 2018 | 0.3 | 2015 | 0.1 | 2017 | 51 | 750 |
| Gabon | Middle | 17,875.76 | 2018 | Upper middle | 2,119,275 | 2018 | 6.3 | 2010 | 0.4 | 2016 | 22 | -- |
| Gambia | West | 2,612.00 | 2018 | Low | 2,280,102 | 2018 | 1.1 | 2011 | 0.1 | 2015 | 4 | 35 |
| Ghana | West | 4,746.68 | 2018 | Lower middle | 29,767,108 | 2018 | 0.9 | 2011 | 0.2 | 2017 | 10 | 640 |
| Guinea | West | 2,504.87 | 2018 | Low | 12,414,318 | 2018 | 0.3 | 2011 | 0.1 | 2016 | 3 | 40 |
| GuineaBissau | West | 1,799.07 | 2018 | Low | 1,874,309 | 2018 | 1 | 2009 | 0.2 | 2015 | 2 | -- |
| Kenya | East | 3,467.56 | 2018 | Lower middle | 51,393,010 | 2018 | 1.4 | 2010 | 0.2 | 2014 | 202 | 586 |
| Lesotho | South | 3,219.46 | 2018 | Lower middle | 2,108,000 | 2018 | 1.3 | 2006 | -- | -- | 5 | -- |
| Liberia | West | 1,308.63 | 2018 | Low | 4,818,977 | 2018 | 0.8 | 2010 | 0.0 | 2015 | -- | -- |
| Libya | North | 20,764.16 | 2018 | Upper middle | 6,678,567 | 2018 | 3.7 | 2014 | 2.2 | 2017 | 330 | 375 |
| Madagascar | East | 1,891.32 | 2018 | Low | 26,262,370,000 | 2018 | 0.2 | 2010 | 0.2 | 2014 | 55 | 200 |
| Malawi | East | 1,311.00 | 2018 | Low | 18,143,310,000 | 2018 | 1.3 | 2011 | 0.0 | 2016 | 6 | 195 |
| Mali | West | 2,316.66 | 2018 | Low | 19,077,690 | 2018 | 0.1 | 2010 | 0.1 | 2016 | 53 | 85 |
| Mauritania | West | 4,150.97 | 2018 | Lower middle | 4,403,319 | 2018 | 0.4 | 2006 | 0.2 | 2017 | 21 | 115 |
| Mauritius | East | 23,750.99 | 2018 | Upper middle | 1,265,300 | 2018 | 3.4 | 2011 | 2.0 | 2015 | 55 | -- |
| Morocco | North | 8,611.71 | 2018 | Lower middle | 36,029,138 | 2018 | 1.1 | 2014 | 0.7 | 2017 | 650 | 1500 |
| Mozambique | East | 1,459.70 | 2018 | Low | 29,495,962 | 2018 | 0.7 | 2011 | 0.1 | 2017 | 60 | 257 |
| Namibia | South | 11,101.83 | 2018 | Upper middle | 2,448,000 | 2018 | 2.7 | 2009 | -- | -- | 60 | 48 |
| Niger | West | 1,063.42 | 2018 | Low | 22,442,948 | 2018 | 0.3 | 2015 | 0.1 | 2014 | 14 | 118 |
| Nigeria | West | 5,990.85 | 2018 | Lower middle | 195,874,740 | 2018 | 0.5 | 2004 | 0.4 | 2013 | 1050 | 250 |
| Rwanda | East | 2,251.56 | 2018 | Low | 12,301,939 | 2018 | 1.6 | 2007 | 0.1 | 2017 | 31 | 454 |
| Sao Tome and Principe | Middle | 3,418.59 | 2018 | Lower middle | 211,028 | 2018 | 2.9 | 2011 | 0.3 | 2015 | -- | -- |
| Senegal | West | 3,782.54 | 2018 | Lower middle | 15,854,360 | 2018 | 0.3 | 2008 | 0.1 | 2016 | 94 | 50 |
| Seychelles | East | 30,557.08 | 2018 | High | 96,762 | 2018 | 3.6 | 2011 | 0.9 | 2016 | 4 |  |
| Sierra Leone | West | 1,601.97 | 2018 | Low | 7,650,154 | 2018 | 0.4 | 2006 | -- | -- | 6 | 72 |
| Somalia | East | -- | -- | Low | 15,008,154 | 2018 | 0.9 | 2014 | 0.0 | 2014 | -- | -- |
| South Africa | South | 13,686.88 | 2018 | Upper middle | 57,780,000 | 2018 | 2.3 | 2010 | 0.9 | 2017 | 8814 | -- |
| South Sudan | East | 1,920.60 | **2014** | Low | 10,975,920 | 2018 | -- | -- | -- | -- | 2 | 40 |
| Sudan | North | 4,767.69 | 2018 | Lower middle | 41,801,533 | 2018 | 0.8 | 2013 | 0.4 | 2015 | 190 | 160 |
| Togo | West | 1,773.90 | 2018 | Low | 7,889,094 | 2018 | 0.7 | 2011 | 0.0 | 2015 | 17 | 203 |
| Tunisia | North | 12,502.82 | 2018 | Lower middle | 11,565,204 | 2018 | 2.3 | 2015 | 1.3 | 2016 | 570 | 0 |
| Uganda | East | 2,038.07 | 2018 | Low | 42,723,139 | 2018 | 0.5 | 2010 | 0.1 | 2015 | 72 | 430 |
| United Republic of Tanzania | East | 3,240.26 | 2018 | Low | 56,318,348 | 2018 | 0.7 | 2010 | 0.0 | 2014 | 50 | 180 |
| Zambia | East | 4,223.91 | 2018 | Lower middle | 17,351,822 | 2018 | 2 | 2010 | 0.1 | 2016 | 79 | 115 |
| Zimbabwe | East | 3,029.79 | 2018 | Lower middle | 14,439,018 | 2018 | 1.7 | 2011 | 0.1 | 2014 | 87 | 193 |

\*Year of data only listed in separate column where there was variation across countries. Otherwise, year of data point is listed under the column name. “—” indicates no data available.

**Appendix 2.**
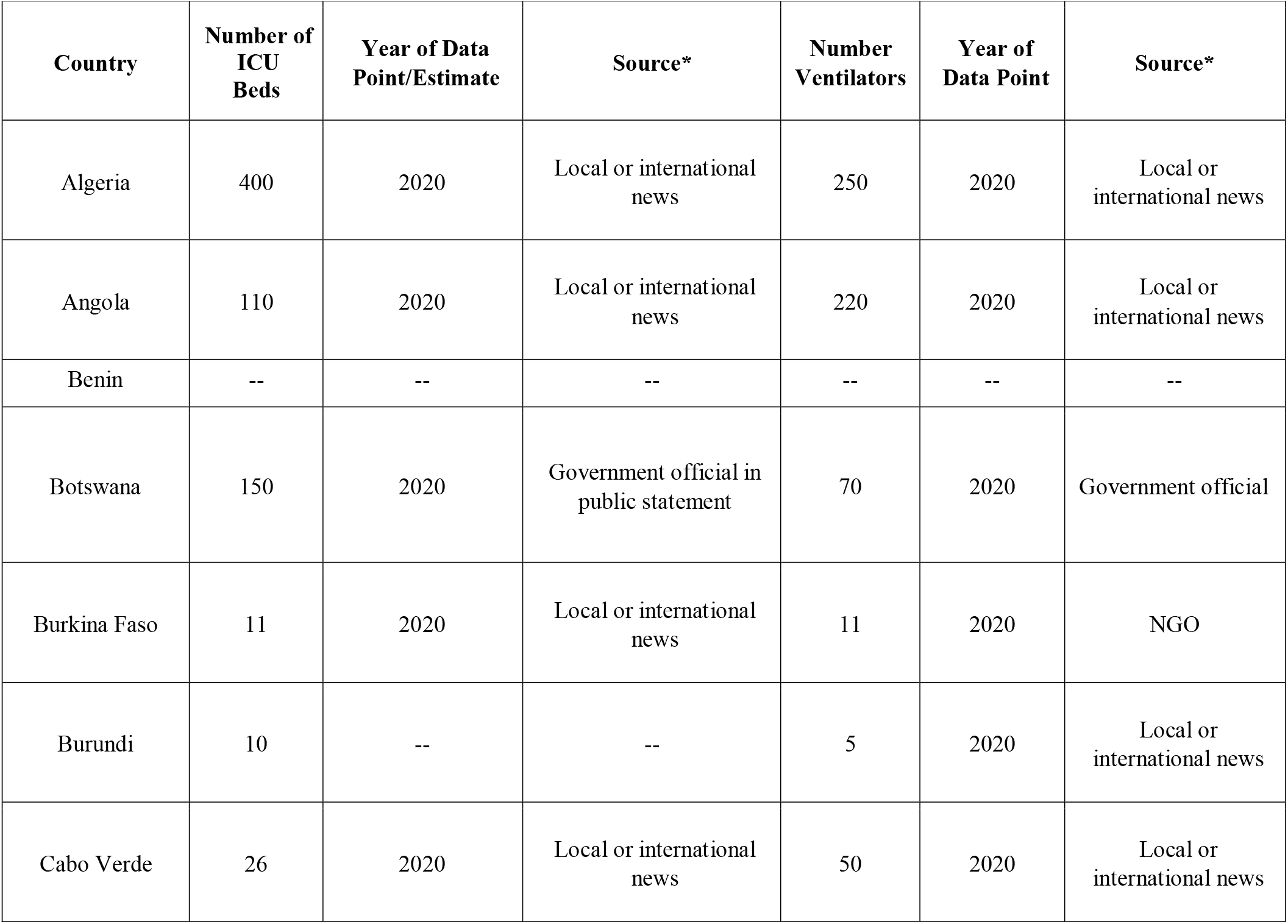
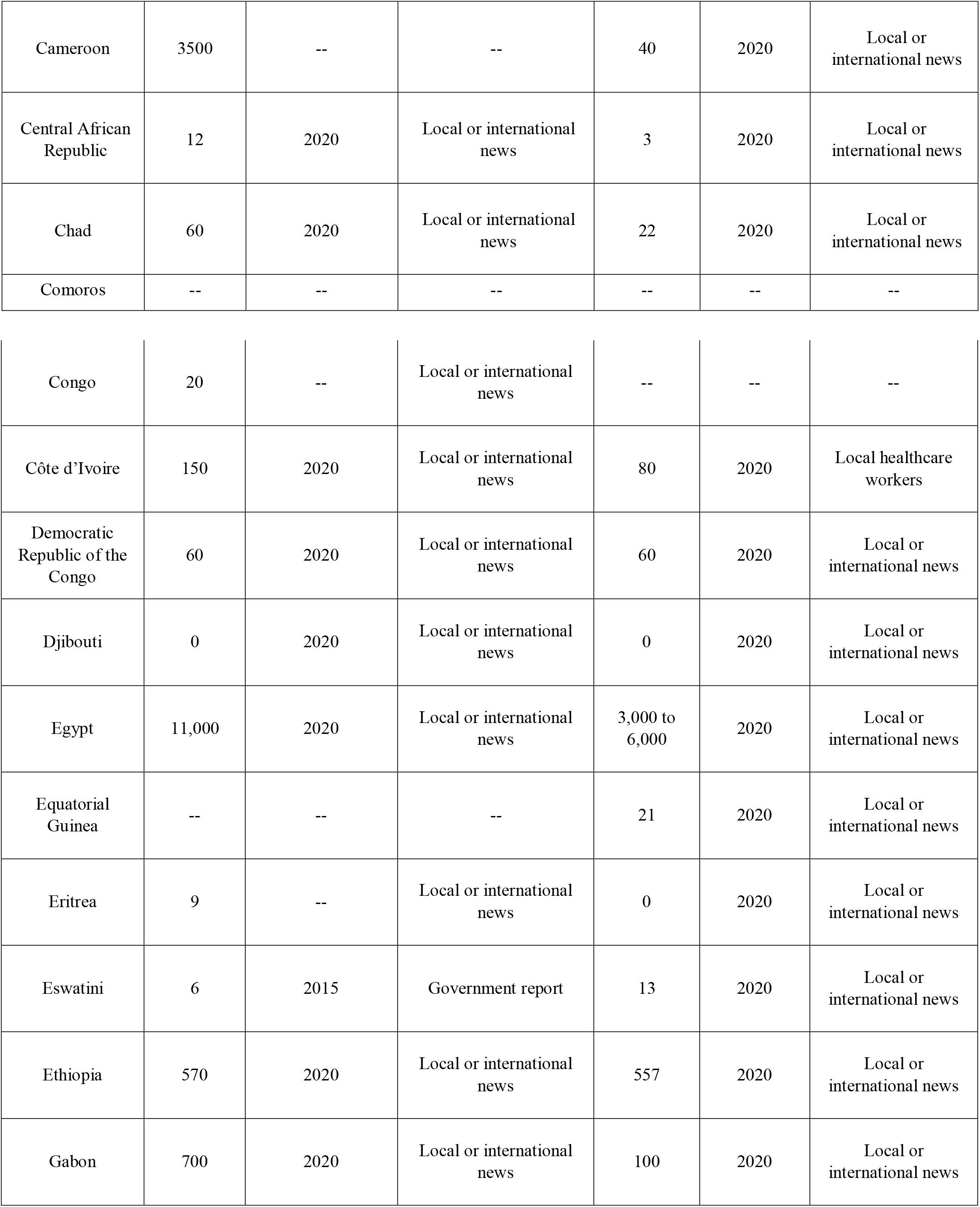
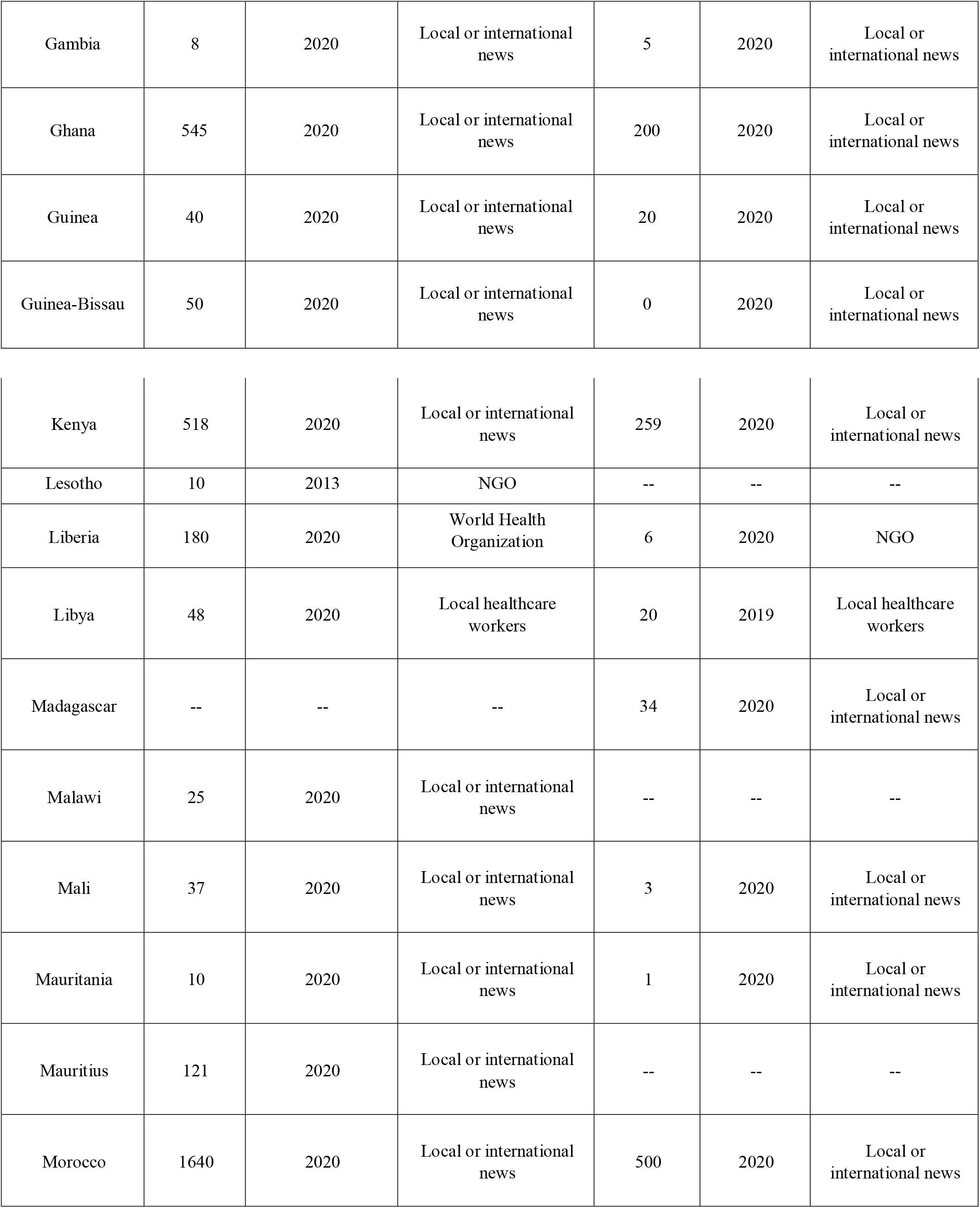
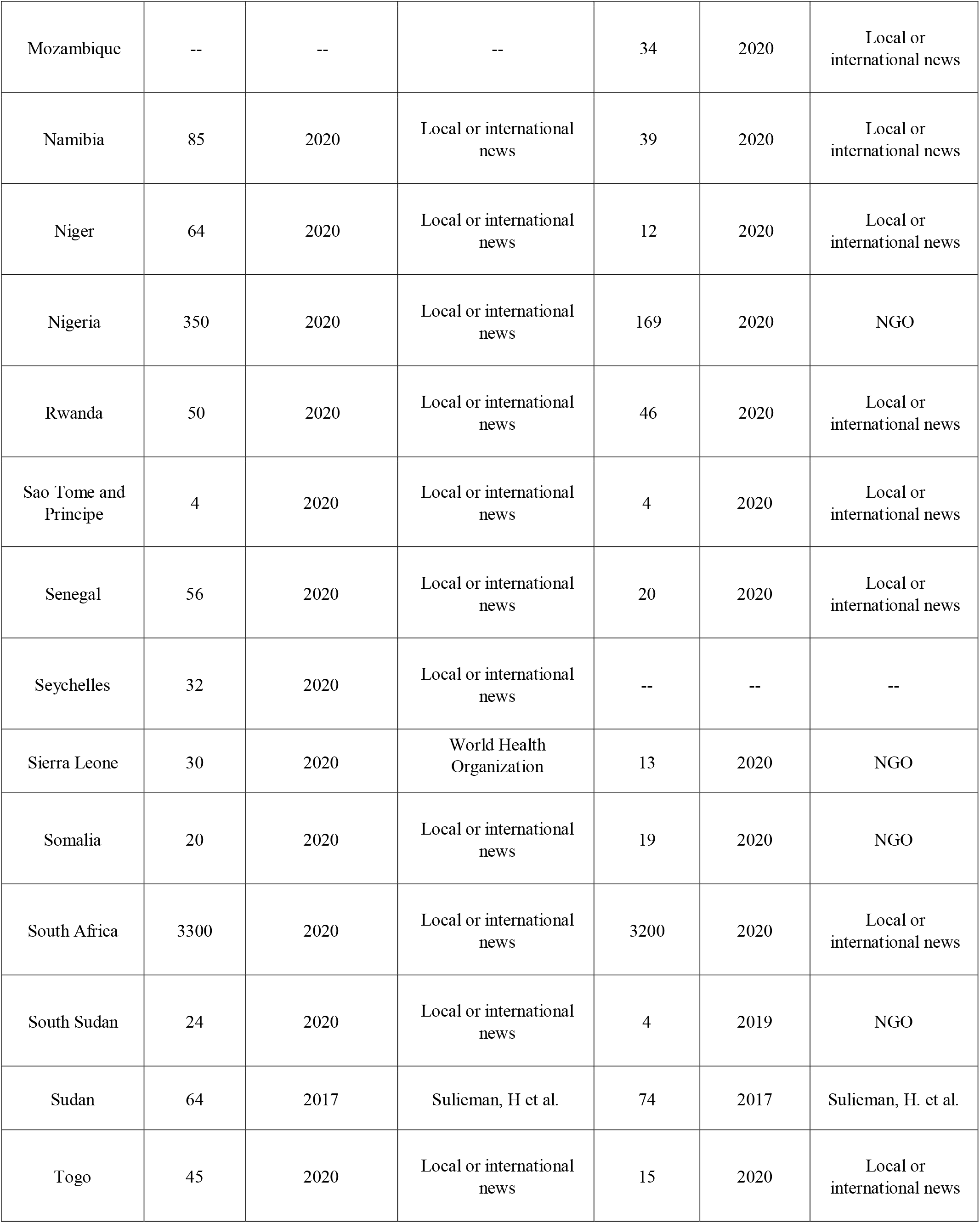
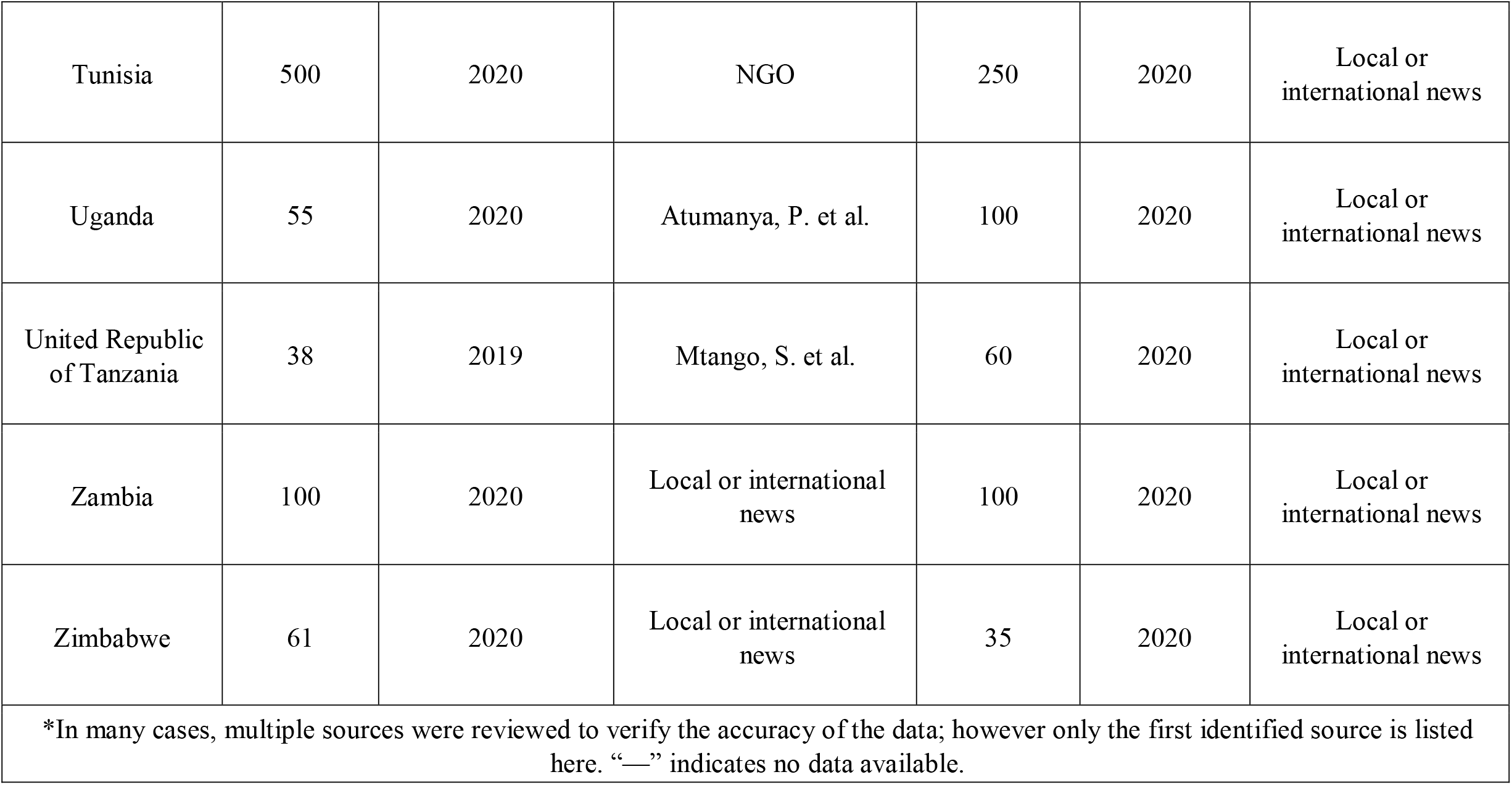
Number of ICU Beds and Ventilators, Year of Estimate and Data Source by Country.

## References

1. World Health Organization. Coronavirus Disease (COVID-19) Situation Report 128. Geneva, Switzerland, 2020.

2. [dataset] World Health Organization. Public health and social measures (PHSMs) database. 2020.

3. Ranney, ML, Griffeth, V, Jha, AK. Critical Supply Shortages — The Need for Ventilators and Personal Protective Equipment during the Covid-19 Pandemic. NJEM 2020, 382: e41.

4. World Health Organization. Shortage of personal protective equipment endangering health workers worldwide, 2020. [accessed 10 May 2020]

5. World Health Organization. Joint External Evaluation (JEE) Mission Reports-Africa Region. 2020.

6. World Health Organization. WHO African Region COVID-19 Readiness Status V2. 2020.

7. Center for Disease Dynamics, Economics & Policy. Modeling COVID-19 in Africa. 2020.

8. Murthy S, Leligdowicz A, Adhikari NKJ. Intensive Care Unit Capacity in Low-Income Countries: A Systematic Review. PLoS One. 2015;10(1):e0116949.

9. Dunser MT. Intensive care medicine in rural sub□Saharan Africa. Anaesthesia 2016; 72(2):181–9. https://doi.org/10.1111/anae.13710.

10. Okafor UV. Challenges in critical care services in Sub-Saharan Africa: Perspectives from Nigeria. Indian J Crit Care Med, 2009; 13(1): 25–7. https://doi.org/10.4103/0972-5229.53112.

11. Kempthorne P, Morriss WW, Mellin-Olsen J, Gore-Booth, J. The WFSA Global Anesthesia Workforce Survey. Anesthesia & Analgesia, 2017; 125(3):981–90.

12. World Federation of Societies of Anaesthesiologists. World Anaesthesiology Workforce Map, https://www.wfsahq.org/workforce-map; 2020 [accessed 04 May 2020].

13. The World Bank. International Comparison Program Database, https://www.worldbank.org/en/programs/icp, 2020 [accessed 04 May 2020].

14. The World Bank. World Development Indicators. 2020.

15. United Nations Population Division. World Population Prospects 2019 – Special Aggregates. 2019.

16. Peterson KFF. Health System Tracker: How prepared is the US to respond to COVID-19 relative to other countries?, https://www.healthsystemtracker.org/chart-collection/how-prepared-is-the-us-to-respond-to-covid-19-relative-to-other-countries/; 2020 [accessed 01 May 2020].

17. Wunsch H, Angus DC, Harrison DA, Collange O, Fowler R, Hoste EA, de Keizer NF, Kersten A, Linde-Zwirble WT, Sandiumenge A, Rowan KM. Variation in critical care services across North America and Western Europe, Crit Care Med, 2008; 36(10): 2787–93. doi:10.1097/CCM.0b013e318186aec8

